# A Case Report Describing a Persistent SARS-CoV-2 Infection Outcomes and Mutations Associated with B-cell Deficiency

**DOI:** 10.64898/2026.02.13.26346281

**Authors:** Rana Mohamed, Alex Shipe, Andrew Lail, Isla Emmen, William Vuyk, Nicholas Minor, Taylor Bradley, Angela Gifford, Nancy Wilson, David H. O’Connor, Jacqueline Garonzik-Wang, Jeannina Smith

## Abstract

**Background:** Immunocompromised (IC) individuals are at increased risk for persistent SARS-CoV-2 infections and can develop new viral mutations and lineages not seen in the community. In this case report, a persistent SARS-CoV-2 infection (330 days) in an IC patient is examined for viral mutations and mutations associated with cryptic lineages.

**Case Presentation:** The patient was followed in a longitudinal study examining persistent SARS-CoV-2 in IC patients. The patient provided stool and nasal swab samples biweekly until 28 days post-enrollment, then monthly, and then quarterly after 12 month post enrollment until the participant was no longer positive for SARS-CoV-2. Staff performed RT-qPCR and viral sequencing on the samples. Viral mutations from the XBK lineage were already present in the initial sample. By the end of the infection period, there were 40 fixed consensus changes from XBK of which two mutations were typical for cryptic lineages. Mutations increased steadily over time, with most mutations fixed by day 253, including the cryptic typical mutations.

**Conclusion:** This case demonstrates the potential for persistent SARS-CoV-2 infections to develop mutations and lineages in IC patients and highlights the need for continued SARS-CoV-2 monitoring and treatment in this vulnerable population.

## Introduction

Immunocompromised (IC) individuals living with B-cell deficiencies have a greater susceptibility to prolonged infections due to their impaired ability to develop antibodies [1,2]. This risk of infection persistence is of particular concern for SARS-CoV-2 infections, as IC patients are at higher risk for severe SARS-CoV-2 infection [3–5] and can become persistent viral shedders leading to increased viral spread [6]. We examined persistent infection in 40 adult IC patients; 39 solid organ transplant (SOT) recipients and one individual with a B-cell deficiency. The data from the SOT cohort were previously published in <u>Vuyk et al</u> [7]. Of 30 SOT recipients, 12 had positive SCV2 RNA at 28 days or more after testing positive; one subject had continual detection out to 54 days after the first positive test. The current report describes the persistent infection and SARS-CoV-2 mutations of the B-cell deficient IC patient who had continued detection of SARS-CoV-2 for 330 days.

## Case Presentation

### Study Timeline

The patient was identified and referred by the PI as a potential study candidate after testing positive for SARS-CoV-2 in December 2023 (d0 days post enrollment - dpe) and self-reporting COVID-19 symptomology for several months. This patient had mantle cell lymphoma in the past, and was treated initially with chemotherapy followed by an autologous stem cell transplant. After 5 years, the mantle cell lymphoma relapsed, and they were treated with lymphodepletion chemotherapy followed by CD 19/20 CAR T infusion. The PI confirmed eligibility based on protocol-specified criteria. Following referral, the research team contacted the patient remotely to discuss participation, and informed consent was obtained. The study was approved by the University of Wisconsin’s IRB.

### Nasal Swab Viral Isolation and RT-qPCR

Nasal Swab samples were self-collected, using nasal swab kits with Universal Transfer Medium (Fisher Scientific, 23600989) and shipped to the processing and storage laboratory overnight. Samples were stored, then batched with other nasal swab samples to isolate on the KingFisher using the Maxwell HT Viral TNA Kit, Custom for the KingFisher (Promega, AX2340). RT-qPCR was performed by the Wisconsin National Primate Center Virology Core, as described in our previous manuscript [7]. Briefly, they used 300 ul of the nasal swab to perform ultra-sensitive quantitative PCR (qPCR) for SARS-CoV-2, analyzing nucleocapsid (N1 and N2) as well as human RNAseP as an internal positive control.

### Stool Sample Viral Isolation and RT-qPCR

Stool samples were self-collected using stool collection kits (Norgen, Thorold, Ontario, Canada). These kits preserve RNA in stool samples for up to 7 days at room temperature. Total nucleic acids (TNA) were isolated from approximately 300 ug of each stool sample using the Promega RSC Fecal Microbiome DNA kit (Promega, Fitchburg, WI, Cat # AS1700). The isolated TNA was tested with qPCR for SARS-CoV-2 nucleocapsid N1 and N2 targets using the GoTaq Enviro Wastewater SARS-CoV-2 System (Promega, Fitchburg, WI, Cat# AM2100) on a LightCycler 96.

### Sequencing

Once we determined that a sample was positive for SARS-CoV-2 and that we had sufficient material for sequencing (Ct<30), we batched samples and used the QIASeq DIRECT SARS-CoV-2 kit (333891) with enhancer (333884) and Region Booster (333897, all Qiagen) to perform overlapping amplicon amplification. Samples were normalized and pooled, then sent to the UW Biotech Center to sequence on the NovaSeq X. Data were analyzed using the oneroof pipeline (github: https://github.com/nrminor/oneroof). Pango lineages were determined using NextClade.

As shown in Figure 1, samples were obtained at day 14 (IC-035-02), day 28 (IC-035-03), and monthly thereafter. After the one-year anniversary, samples were collected quarterly to reduce participant burden. Once the participant tested negative for SARS-CoV-2 in both the nasal swab and stool samples, they were removed from the study. Our last samples from this participant were collected at timepoint IC-035-15, at which time both the stool and nasal swab samples tested negative. We have not included analysis of stool sample sequencing in this study, because the quality of the sequence from stool was not sufficient.

**Figure 1.**
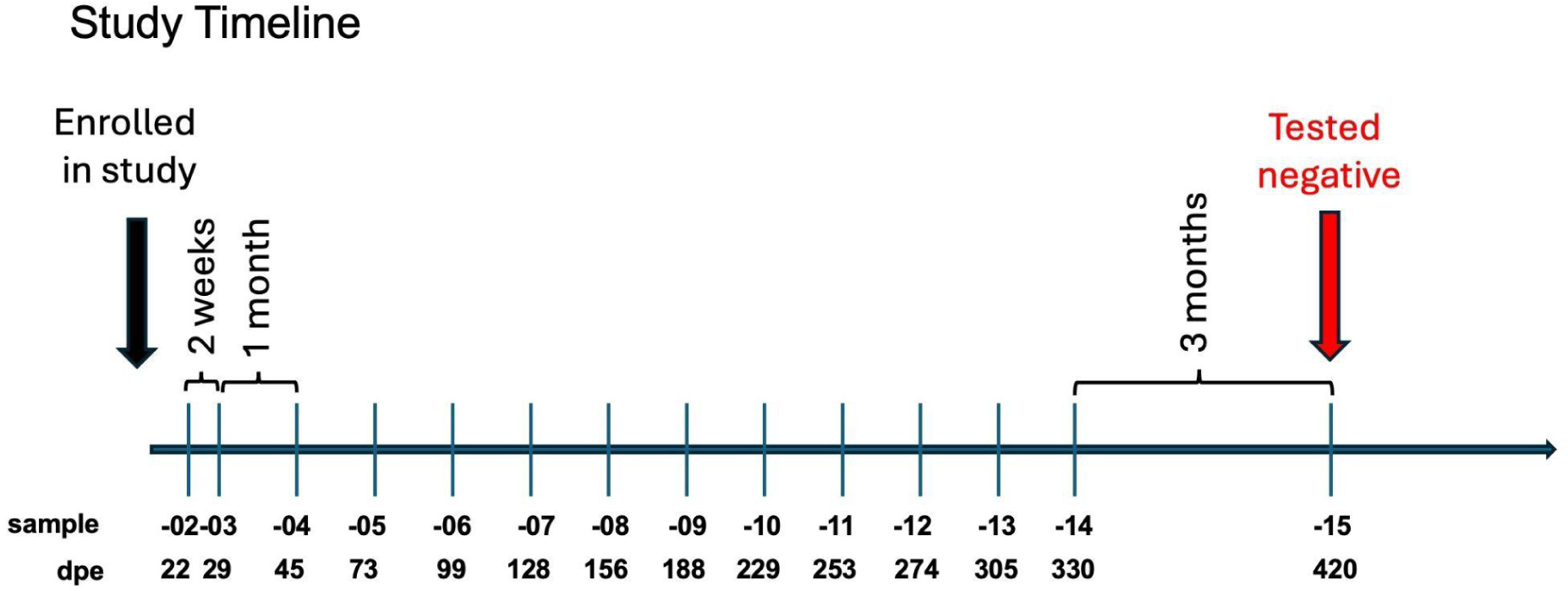
**Timeline for study**: Participants were identified by study physicians and then enrolled by Department of Surgery Clinical Research personnel. Typically, a sample was obtained within one week of enrollment (sample-01), then at weeks 2 and 4, and monthly thereafter. After the first year on-study, samples were obtained quarterly. The maximum enrollment was 2 years. For the current patient, the first sample was delayed longer than usual, so it became sample-02, and there was no sample-01.

### Mutations from the reference baseline isolate of XBK were already present at the earliest timepoint

Although the patient was enrolled in our study in early 2024, sequencing at the first timepoint revealed that the XBK lineage caused the infection. XBK is a recombinant between BA.5.2 and CJ.1, first isolated in Denmark in October 2022. As no GenBank accession is specified in the original Pango designation for XBK, we used the earliest available GenBank genome assigned to lineage XBK (OY837126; Denmark, collected 14 Oct 2022) as a representative reference sequence for comparative analyses. The peak of circulation for this virus was between November 2022 and April 2023 (Figure 2). Therefore, it is likely that the patient was infected between 8 months and 1 year before enrollment into the study. In all, 472 sequences have been uploaded to GenBank, of which 82 were from the USA, 47 were collected in WI, and 34 were from this participant.

**Figure 2.**
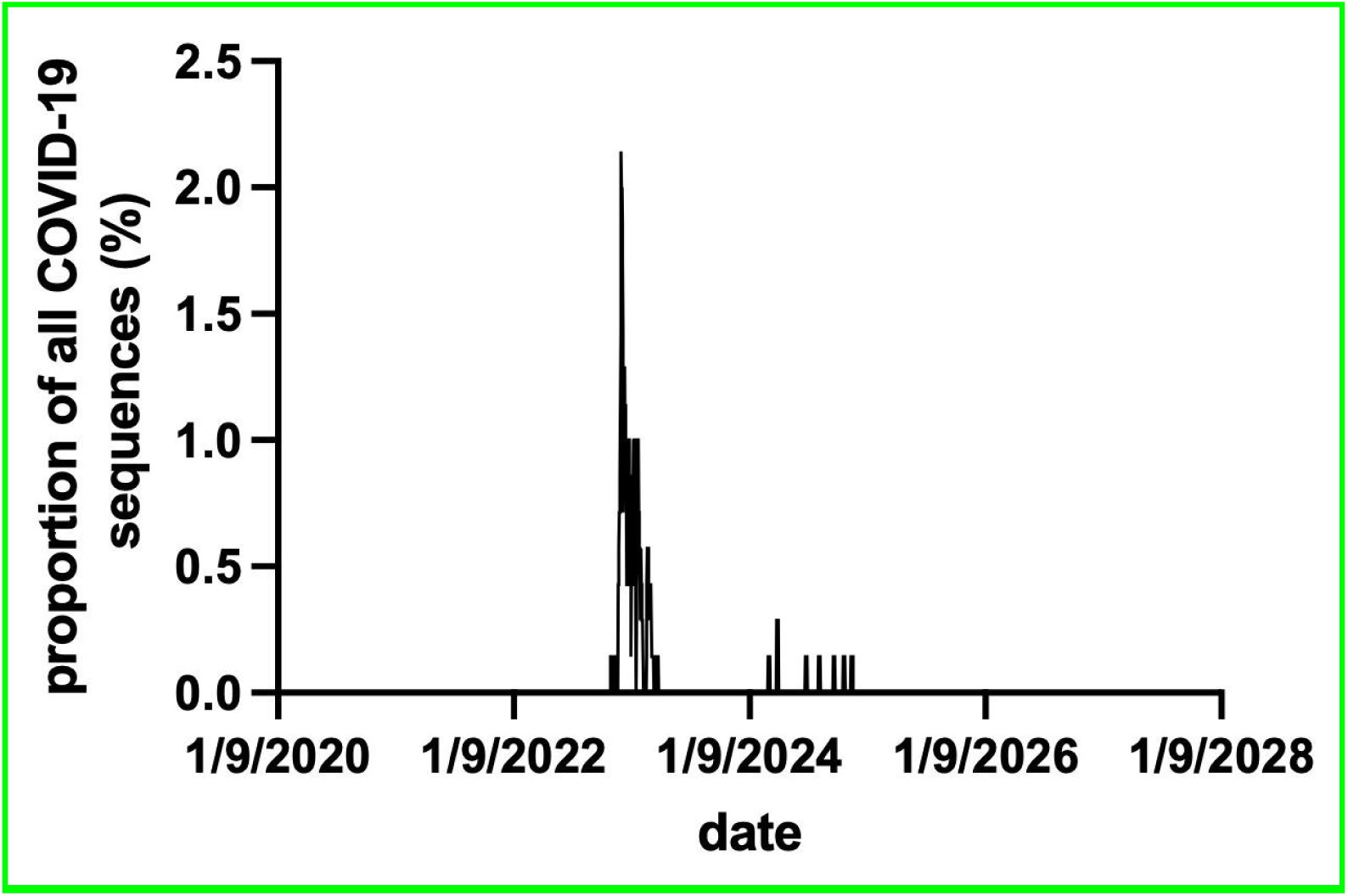
XBK was primarily in circulation between November 2022 and April 2023, according to <u>CovSpectrum.com</u>. The small spikes in 2024 are the patient’s samples.

At the earliest timepoint, 22 dpe, there were already 8 consensus mutations when comparing to the baseline reference XBK sequence (Table 1, Table 2), potentially reflecting ongoing mutation between the time the participant was infected and the first samples. We compared 19 other XBK sequences from the USA that were the most recent entries in GenBank, excluding our IC-035 uploaded sequences to IC-035-NS. We found only one sequence that was closer to our earliest sequence, with 6 mutational differences, a sequence from IL on 1/28/23, nearly a year before. All of the other sequences were more disparate from IC-035-02 (Supplemental Table 1). Therefore, we felt comfortable using the baseline reference, as we did not appear to be biasing the data.

**Table 1.** The reference baseline isolate of the XBK lineage had 65 mutations from Wuhan1. This is a list of changes found in the original XBK sequence (OY837126) compared to Wuhan-1. When analyzing consensus mutations over time, these mutations were removed so that we could determine which mutations were due to changes in XBK attributable to evolution while the patient was infected.

| mutation | mutation |
| --- | --- |
| orf1ab:p.Ser135Arg | ORF1a:S135R |
| orf1ab:p.Thr842Ile | ORF1a:T842I |
| orf1ab:p.Ser1221Leu | ORF1a:S1221L |
| orf1ab:p.Gly1307Ser | ORF1a:G1307S |
| orf1ab:p.Pro1640Ser | ORF1a:P1640S |
| orf1ab:p.Leu3027Phe | ORF1a:T3090I |
| orf1ab:p.Thr3090Ile | ORF1a:L3201F |
| orf1ab:p.Leu3201Phe | ORF1a:T3255I |
| orf1ab:p.Thr3255Ile | ORF1a:P3395H |
| orf1ab:p.Pro3395His | ORF1a:del3675/3677 |
| orf1ab:p.Asn4060Ser | ORF1a:N4060S |
| orf1ab:p.Pro4715Leu | ORF1b:P314L |
| orf1ab:p.Gly5063Ser | ORF1b:G662S |
| orf1ab:p.Arg5716Cys | ORF1b:R1315C |
| orf1ab:p.Ile5967Val | ORF1b:I1566V |
| orf1ab:p.Val6107Ile | ORF1b:V1706I |
| orf1ab:p.Thr6564Ile | ORF1b:T2163I |
| S:p.Thr19Ile | S:T19I |
| S:p.Ala27Ser | S:A27S |
| S:p.Gly142Asp | S:G142D |
| S:p.Lys147Glu | S:K147E |
| S:p.Trp152Arg | S:W152R |
| S:p.Phe157Leu | S:F157L |
| S:p.Ile210Val | S:I210V |
| S:p.Val213Gly | S:V213G |
| S:p.Gly257Ser | S:G257S |
| S:p.Gly339His | S:G339H |
| S:p.Arg346Thr | S:R346T |
| S:p.Ser371Phe | S:S371F |
| S:p.Ser373Pro | S:S373P |
| S:p.Ser375Phe | S:S375F |
| S:p.Thr376Ala | S:T376A |
| S:p.Asp405Asn | S:D405N |
| S:p.Arg408Ser | S:R408S |
| S:p.Lys417Asn | S:K417N |
| S:p.Asn440Lys | S:N440K |
| S:p.Gly446Ser | S:G446S |
| S:p.Asn460Lys | S:N460K |
| S:p.Ser477Asn | S:S477N |
| S:p.Thr478Lys | S:T478K |
| S:p.Glu484Ala | S:E484A |
| S:p.Phe483Pro | S:F486P |
| S:p.Phe490Ser | S:F490S |
| S:p.Gln498Arg | S:Q498R |
| S:p.Asn501Tyr | S:N501Y |
| S:p.Tyr505His | S:Y505H |
| S:p.Asp614Gly | S:D614G |
| S:p.His655Tyr | S:H655Y |
| S:p.Asn679Lys | S:N679K |
| S:p.Pro681His | S:P681H |
| S:p.Asn764Lys | S:N764K |
| S:p.Asp796Tyr | S:D796Y |
| S:p.Gln954His | S:Q954H |
| S:p.Asn969Lys | S:N969K |
| ORF3a:p.Thr223Ile | ORF3a:T223I |
| E:p.Thr9Ile | E:T9I |
| E:p.Thr11Ala | E:T11A |
| M:p.Gln19Glu | M:Q19E |
| M:p.Ala63Thr | M:A63T |
| ORF6:p.Asp61Leu | ORF6:D61L |
| N:p.Pro13Leu | N:P13L |
| N:p.Arg203Lys | N:R203K |
| N:p.Gly204Arg | N:G204R |
| N:p.Ser413Arg | N:S413R |
| ORF9b:p.Pro10Ser | ORF9b:P10S |

**Table 2.**
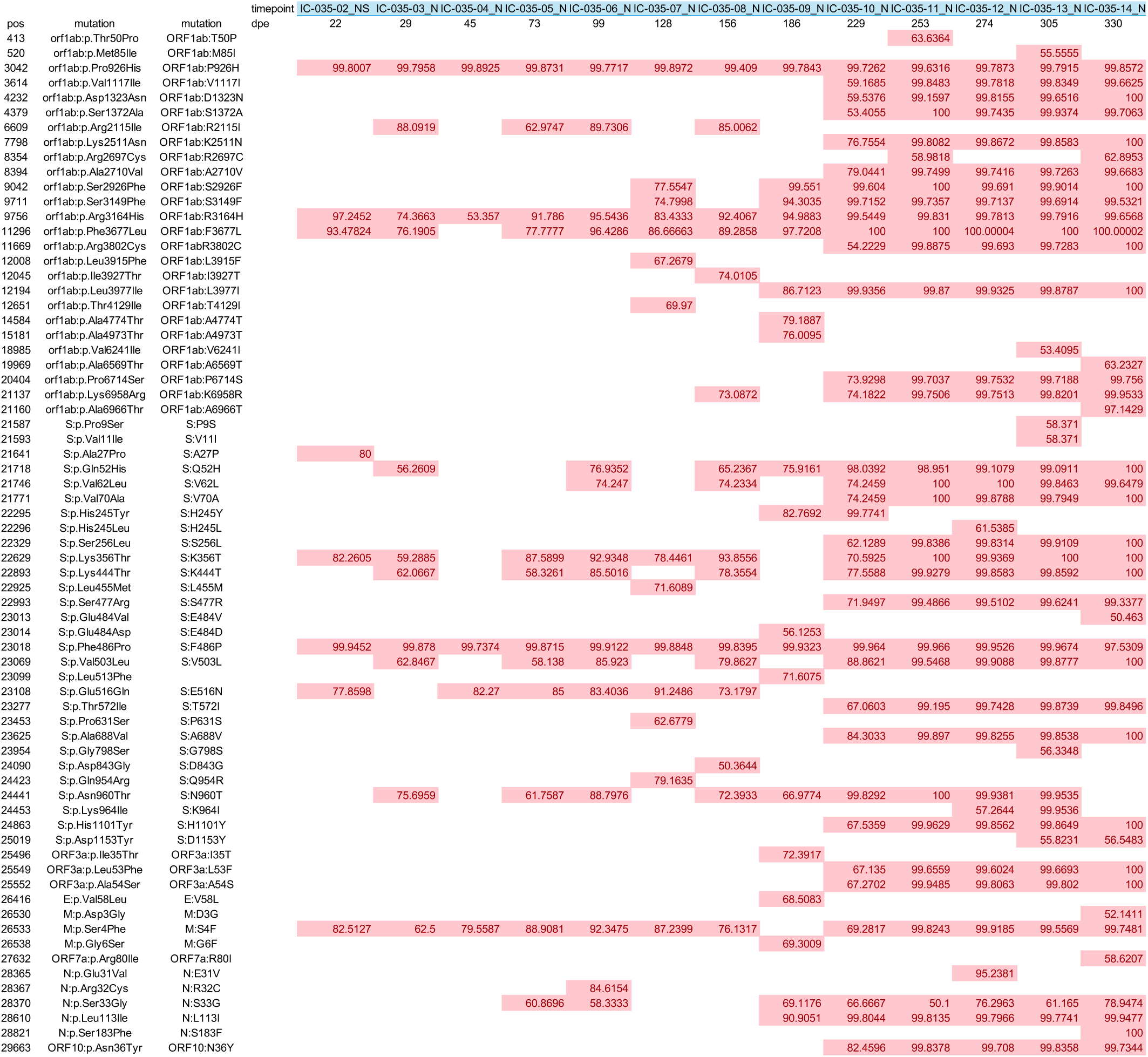
Consensus mutations over time. This table shows the frequencies of mutations that were observed in nasal swab sequences from this patient over time. We have highlighted them in red so that it is easier to observe the increase in the number of mutations over time visually. We took the individual variant.tsv files and concatenated them, then filtered all of the changes by missense mutations. These data were converted into a pivot table and sorted using the genomic position, using a script written by Claude AI. We determined which mutations were at any timepoint at or above consensus by filtering on frequencies greater than 50%. Then we removed all of the mutations present in the reference baseline isolate of XBK compared to Wuhan-1, so that we can look at only variants that developed after the patient was infected.

### Increasing mutations over time

With prolonged infection, we expected that SARS-CoV-2 would continue accruing within-host variants. We plotted the number of consensus mutations over time, and found a strong positive correlation between days post enrollment (dpe) and total consensus mutation burden (Pearson’s R^2^=0.92, p<0.0001). This was consistent with progressive mutation accumulation over time. (Figure 3, black circles/line, Table 2) Overlaying mutation burden with a sweep index (Figure 3, blue bars, Table 3), we showed that the number of variants newly reaching near-fixation (≥95% frequency) demonstrated that fixation events occurred episodically rather than continuously (Table 4). While the total variant burden increased gradually, large clusters of variants crossed the fixation threshold simultaneously, most notably at timepoint d253, with 22 variants reaching ≥95% frequency at this timepoint. Importantly, these fixation events were not accompanied by abrupt increases in total variant count, suggesting that multiple variants swept together as linked sets. This pattern was consistent with episodic clonal or haplotype replacement within the viral population, rather than independent, gradual fixation of individual variants.

**Figure 3.**
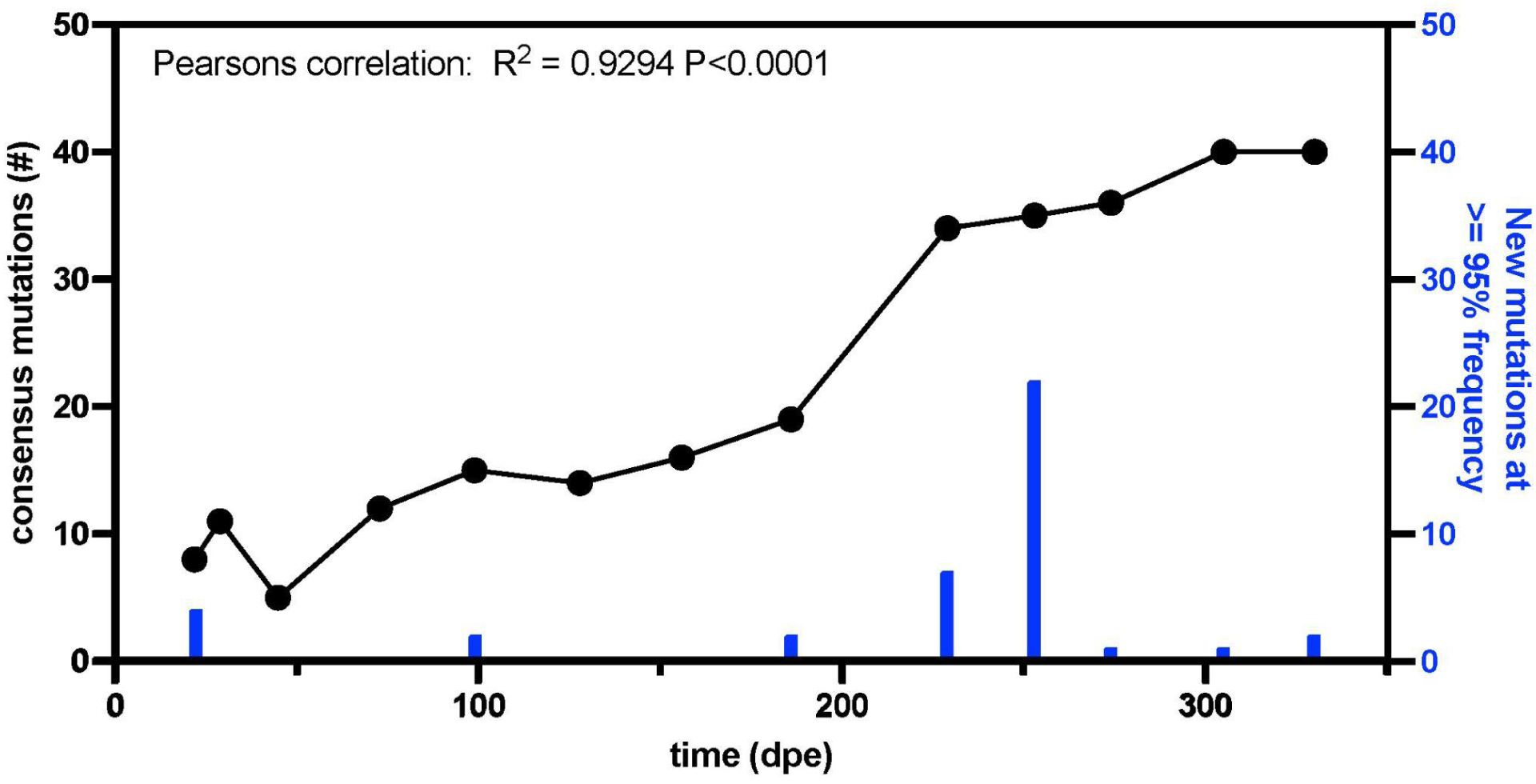
Variants that were at 50% frequency or more, that were not present in the reference baseline isolate of the XBK lineage, increased over time in a largely linear fashion. Total variant burden (black circles and line) and sweep index (blue bars), defined as the number of variants newly crossing ≥95% frequency at each timepoint, are shown over time post-enrollment. The variant burden increases gradually and approximately linearly, whereas fixation events occur in discrete bursts. The largest sweep at timepoint dpe 253 involves simultaneous fixation of multiple variants without a corresponding jump in total variant burden, consistent with clonal or haplotype-level replacement rather than independent fixation of individual mutations.

**Table 3.** Sweep data. . To quantify the timing and magnitude of selective sweeps, we defined a sweep index as the number of variants whose population frequency crossed a dominance threshold, in this case 95%, between consecutive timepoints. Fixation dynamics are dominated by a single major sweep at day 253, during which over twenty variants reach near-fixation simultaneously.

| Timepoint | New $\geq 95\%$ |
| --- | --- |
| 22 | 4 |
| 29 | 0 |
| 45 | 0 |
| 73 | 0 |
| 99 | 2 |
| 128 | 0 |
| 156 | 0 |
| 186 | 2 |
| 229 | 7 |
| **253** | **22** |
| 274 | 1 |
| 305 | 1 |
| 330 | 2 |

**Table 4.**
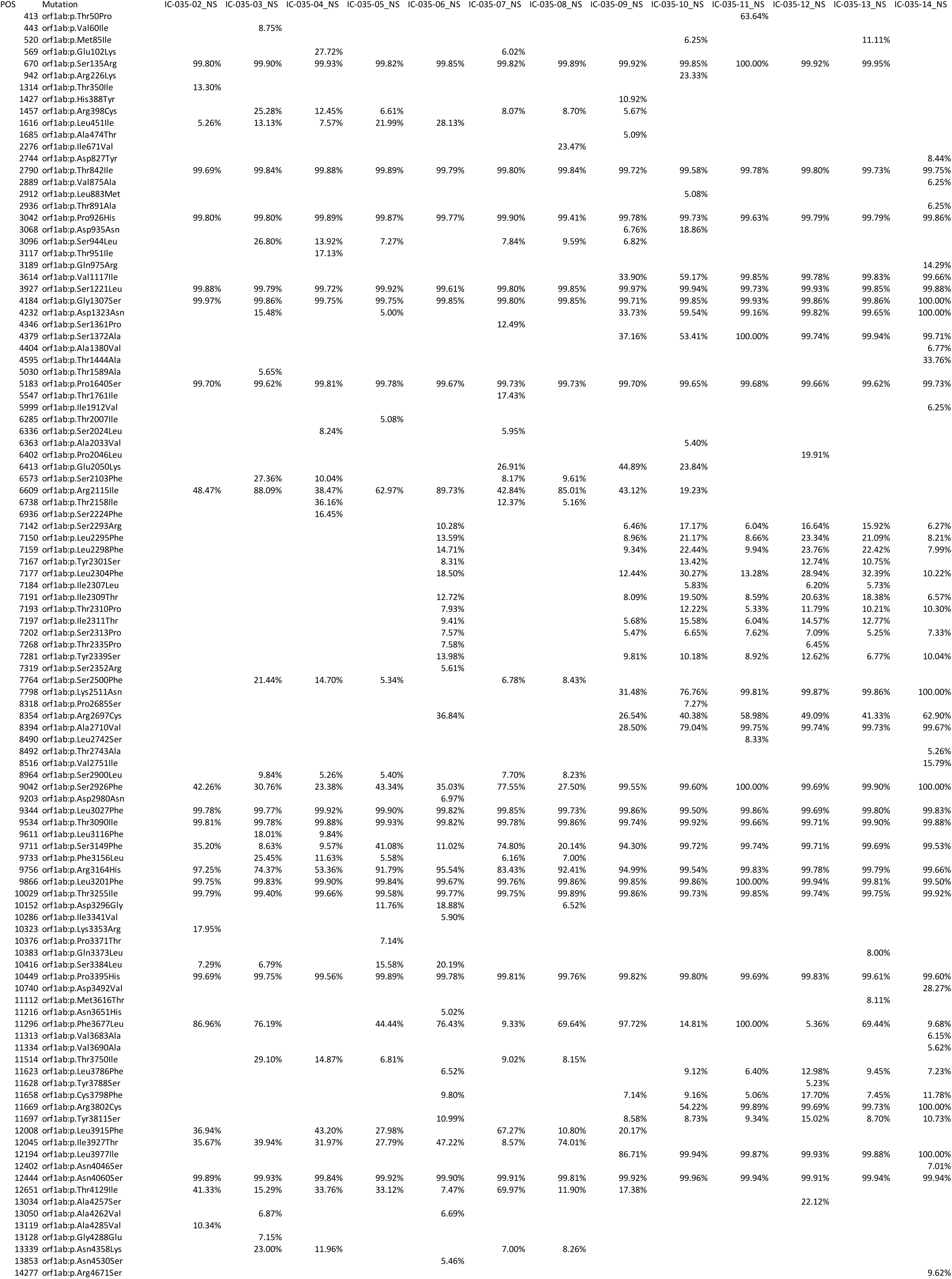
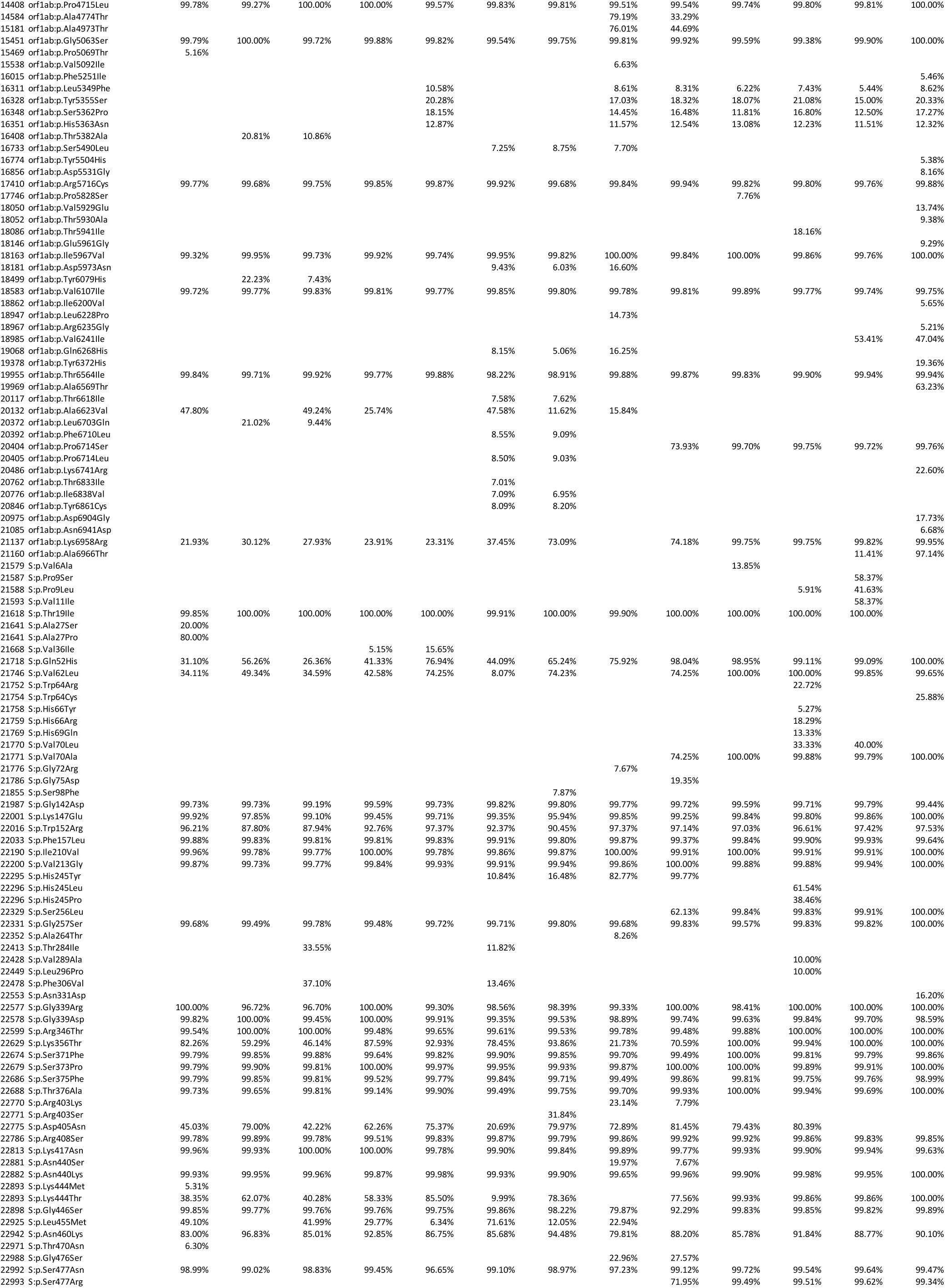
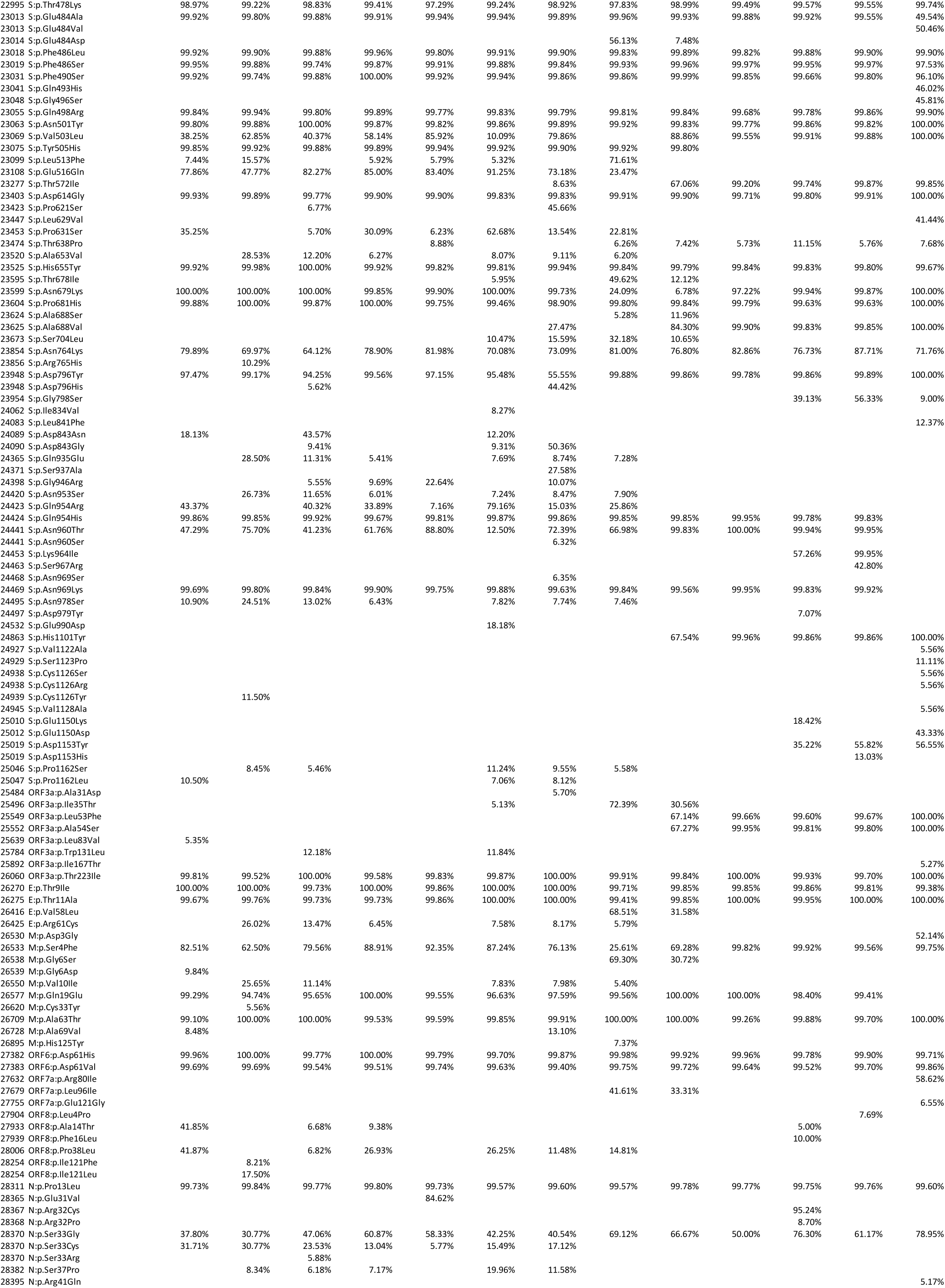
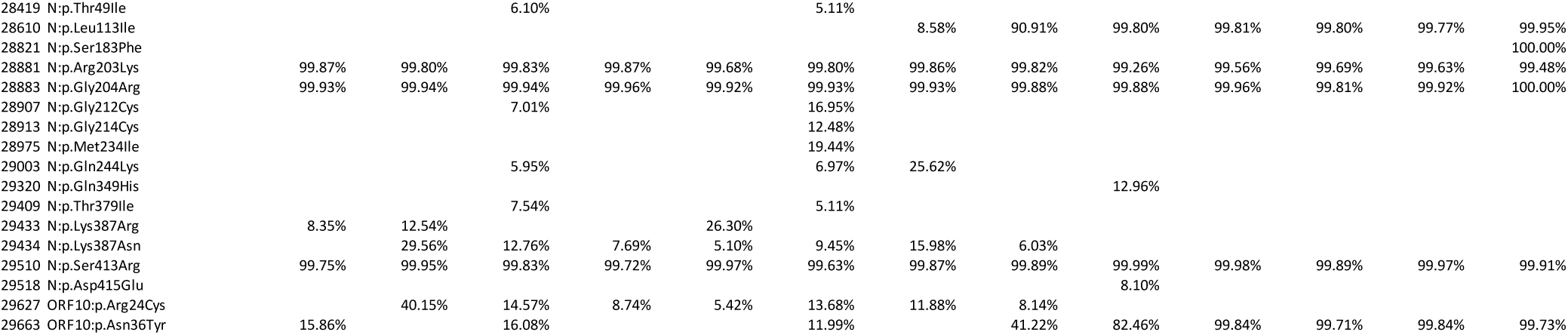
Pivot table with all variant frequency data. We took the individual variant.tsv files and concatenated them, then filtered all of the changes by missense mutations. These data were converted into a pivot table and sorted using the genomic position, using a script written by Claude AI. The script is in our data portal. Note that at some timepoints, oneroof outputs two different variants, both at >90% frequency. For these cases, the variants listed are incorrect because the actual variant present is due to a codon that has two changes and results in a totally different amino acid residue than either of those listed. This occurred three times: ORF6:D61L, S:G339H, and S:F486P.

Cryptic lineages have been hypothesized to emerge in immunocompromised patients with prolonged viral replication [8–10]. Consistent with this, we observed the emergence of two mutations often seen in cryptic lineages, S:K444T and S:T572I at d229, becoming fixed at d253.

## Discussion

In this case report, the patient tested positive for SARS-CoV-2 in either nasal swabs or stool for almost a year and reported experiencing COVID-like symptoms for several months prior to enrollment. Based on the XBK variant we sequenced, and the time frame when that variant was most prevalent in the community, it is likely that this immunocompromised participant had been infected for at least several months before enrolling in the study. Although new variants arose throughout the observation period, fixation occurred episodically, with large clusters of mutations swept to near-fixation simultaneously. This pattern is consistent with clonal (haplotype-level) replacement rather than independent, gradual fixation of individual mutations. These new mutations included two that are often seen in cryptic lineages.

Persistent SARS-CoV-2 infections endanger the individual and, more broadly, the population by sustaining the potential for transmission to others. According to the CDC, it is estimated that from October 2024 to September 2025, there have been between 13.8-20.3 million infections of SARS-CoV-2 and 44,000-63,000 deaths due to SARS-CoV-2 in the United States [11]. In the general population, 6% of SARS-CoV-2 infections have been shown to have viral shedding after 30 days since onset, and it is estimated that between 0.1-0.5% of individuals who contract SARS-CoV-2 infection become persistently infected [12]. *Swank, Z., et. al.,* found that 40% of individuals with long COVID symptoms in the post-acute phase tested antigen-positive and 20% of all individuals had persistent antigen production between 1-month and 14-months post-infection[13].

The true prevalence of SARS-CoV-2 infections, viral shedding, and persistent cases in the population remains uncertain. However, immunocompromised individuals are at increased risk for severe and prolonged SARS-CoV-2 infection due to impaired immune-mediated viral clearance [3–5]. In this population, COVID-19 may follow a persistent or relapsing course with extended viral shedding and increased morbidity. Population-level data from the INFORM study demonstrate that immunocompromised patients continue to account for a disproportionate share of COVID-19–related hospitalizations and deaths, underscoring their ongoing vulnerability [14].

The viral mutations noted in persistent SARS-CoV-2 infections in IC individuals are relevant for treatment and management of SARS-CoV-2. Our patient exhibited two viral mutations with changes often found in cryptic lineages. Leveraging novel cellular therapies may help to curtail the length of infections and mutations seen in these cases. Direct-acting antiviral treatment has also shown promise in the treatment of persistent SARS-CoV-2 in mouse models, but clinical trials are needed to better understand patient outcomes [15]. While people experiencing long COVID may utilize longer duration Paxlovid [16,17], this is difficult in a person on transplant immunosuppression because ritonavir is a very strong CYP3A4 inhibitor and calcineurin inhibitors are metabolized by CYP3A4, potentially resulting in toxic blood levels of these medications [18–20].

Our patient received several SARS-CoV-2–directed therapies before enrollment in our study. Two years before enrollment, after testing positive for COVID-19, the patient received a single intravenous infusion of casirivimab/imdevimab (REGEN-COV). The following year, the patient was given tixagevimab (300 mg) and cilgavimab (300 mg) (Evusheld) as a one-time injection for pre-exposure prophylaxis.

In March 2023, the patient was treated with remdesivir for 4 days in combination with dexamethasone (6 mg daily for 10 days), a regimen shown to be effective against the Omicron variant and which remains FDA-approved for the treatment of COVID-19 [11,12]. However, while remdesivir plus dexamethasone is effective as antiviral therapy, it does not provide the same duration of protection as monoclonal antibody therapy.

As of 2025, neither casirivimab/imdevimab nor tixagevimab/cilgavimab is used for SARS-CoV-2 treatment due to reduced activity against currently circulating variants. Nine months after the March 2023 treatment, the patient was enrolled in our study.

## Conclusion

Persistent SARS-CoV-2 infections in immunocompromised individuals continue to be an area of concern for the development of SARS-CoV-2 variants. This case report of an immunocompromised patient living with a B-Cell deficiency who tested positive for SARS-CoV-2 for nearly a year highlights the time-dependent nature of SARS-CoV-2 viral mutation and the potential for new cryptic lineages to arise from long-term infections. Continued efforts to develop anti-viral therapies that can provide adequate protection from SARS-CoV-2 infection persistence could improve outcomes for immunocompromised patients and slow the development of new lineages over time.

## Data availability

Data are available at the following portal: https://dholk.primate.wisc.edu/dho/public/manuscripts/published/Persistent%20SARS-CoV-2%[…]with%20B-Cell%20Deficiency/project-begin.view?wizard=true

## Acknowledgements

We would like to acknowledge Marc Johnson, an expert in cryptic lineages of SARS-CoV-2, for his contributions to interpreting our cryptic lineage results.

---

**Table S1.**
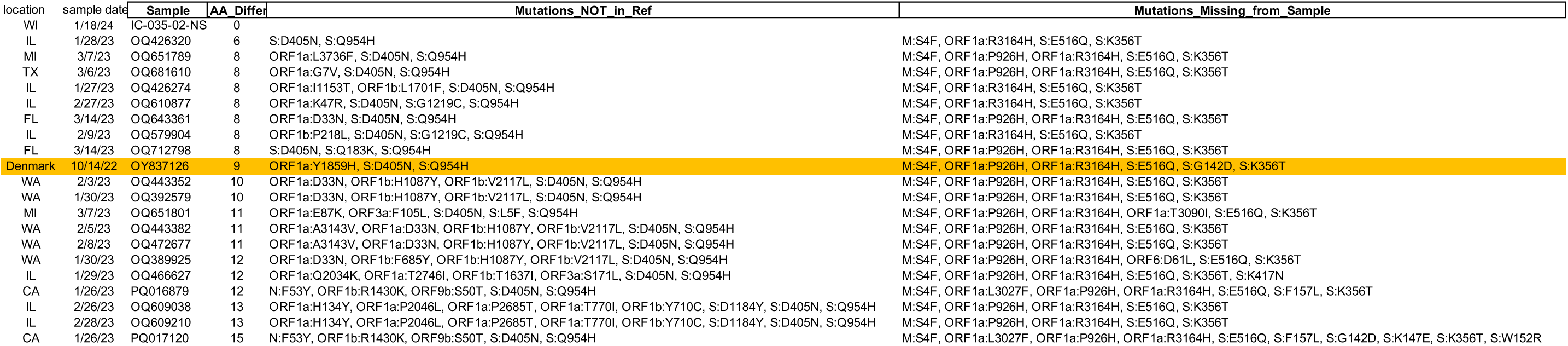
Similarity Analysis between IC-035-02 and XBK sequences uploaded to GenBank that were nearer in time and distance to IC-035 than the baseline reference sequence we chose. We downloaded a listing of all of the XBK sequences that had been uploaded to GenBank. The earliest, OY837126, was chosen as the baseline lineage upon which we based our analysis of consensus mutation changes over time. We wondered if we were biasing the data by this choice. Therefore, we filtered the list by samples taken in the USA, and looked for the 19 most recent samples that were not samples we uploaded. We input these 19, along with OY837126 and IC-035-02 into Nextclade using the Wuhan strain as the comparator sequence. The output nextclade.tsv file was analyzed in python (Compare_XBK.ipynb - in the data portal), the output is a similarity analysis in which each sequence is compared to IC-035-02, then ranked by the number and kind of mutations. Mutations_NOT_in_Ref are mutations present in the comparator sequences that are not present in IC-035-02, but are present in those sequences. Mutation_Missing_from_sample are mutations present in IC-035-02, but not in the comparator sequence. The baseline reference sequence is highlighted in Orange and is in the middle of all the sequences expected to be most like IC-035-02. Therefore, we do not feel that our choice of this sequence as the baseline reference is biased.

| location | sample date | Sample | AA Differ | Mutations_NOT_in_Ref | Mutations_Missing_from_Sample |
| --- | --- | --- | --- | --- | --- |
| WI | 1/18/24 | IC-035-02-NS | 0 |  |  |
| IL | 1/28/23 | OQ426320 | 6 | S:D405N, S:Q954H | M:S4F, ORF1a:R3164H, S:E516Q, S:K356T |
| MI | 3/7/23 | OQ651789 | 8 | ORF1a:L3736F, S:D405N, S:Q954H | M:S4F, ORF1a:P926H, ORF1a:R3164H, S:E516Q, S:K356T |
| TX | 3/6/23 | OQ681610 | 8 | ORF1a:G7V, S:D405N, S:Q954H | M:S4F, ORF1a:P926H, ORF1a:R3164H, S:E516Q, S:K356T |
| IL | 1/27/23 | OQ426274 | 8 | ORF1a:I1153T, ORF1b:L1701F, S:D405N, S:Q954H | M:S4F, ORF1a:R3164H, S:E516Q, S:K356T |
| IL | 2/27/23 | OQ610877 | 8 | ORF1a:K47R, S:D405N, S:G1219C, S:Q954H | M:S4F, ORF1a:R3164H, S:E516Q, S:K356T |
| FL | 3/14/23 | OQ643361 | 8 | ORF1a:D33N, S:D405N, S:Q954H | M:S4F, ORF1a:P926H, ORF1a:R3164H, S:E516Q, S:K356T |
| IL | 2/9/23 | OQ579904 | 8 | ORF1b:P218L, S:D405N, S:G1219C, S:Q954H | M:S4F, ORF1a:R3164H, S:E516Q, S:K356T |
| FL | 3/14/23 | OQ712798 | 8 | S:D405N, S:Q183K, S:Q954H | M:S4F, ORF1a:P926H, ORF1a:R3164H, S:E516Q, S:K356T |
| Denmark | 10/14/22 | OY837126 | 9 | ORF1a:Y1859H, S:D405N, S:Q954H | M:S4F, ORF1a:P926H, ORF1a:R3164H, S:E516Q, S:G142D, S:K356T |
| WA | 2/3/23 | OQ443352 | 10 | ORF1a:D33N, ORF1b:H1087Y, ORF1b:V2117L, S:D405N, S:Q954H | M:S4F, ORF1a:P926H, ORF1a:R3164H, S:E516Q, S:K356T |
| WA | 1/30/23 | OQ392579 | 10 | ORF1a:D33N, ORF1b:H1087Y, ORF1b:V2117L, S:D405N, S:Q954H | M:S4F, ORF1a:P926H, ORF1a:R3164H, S:E516Q, S:K356T |
| MI | 3/7/23 | OQ651801 | 11 | ORF1a:E87K, ORF3a:F105L, S:D405N, S:L5F, S:Q954H | M:S4F, ORF1a:P926H, ORF1a:R3164H, ORF1a:T3090I, S:E516Q, S:K356T |
| WA | 2/5/23 | OQ443382 | 11 | ORF1a:A3143V, ORF1a:D33N, ORF1b:H1087Y, ORF1b:V2117L, S:D405N, S:Q954H | M:S4F, ORF1a:P926H, ORF1a:R3164H, S:E516Q, S:K356T |
| WA | 2/8/23 | OQ472677 | 11 | ORF1a:A3143V, ORF1a:D33N, ORF1b:H1087Y, ORF1b:V2117L, S:D405N, S:Q954H | M:S4F, ORF1a:P926H, ORF1a:R3164H, S:E516Q, S:K356T |
| WA | 1/30/23 | OQ389925 | 12 | ORF1a:D33N, ORF1b:F685Y, ORF1b:H1087Y, ORF1b:V2117L, S:D405N, S:Q954H | M:S4F, ORF1a:P926H, ORF1a:R3164H, ORF6:D61L, S:E516Q, S:K356T |
| IL | 1/29/23 | OQ466627 | 12 | ORF1a:Q2034K, ORF1a:T2746I, ORF1b:T1637I, ORF3a:S171L, S:D405N, S:Q954H | M:S4F, ORF1a:P926H, ORF1a:R3164H, S:E516Q, S:K356T, S:K417N |
| CA | 1/26/23 | PQ016879 | 12 | N:F53Y, ORF1b:R1430K, ORF9b:S50T, S:D405N, S:Q954H | M:S4F, ORF1a:L3027F, ORF1a:P926H, ORF1a:R3164H, S:E516Q, S:F157L, S:K356T |
| IL | 2/26/23 | OQ609038 | 13 | ORF1a:H134Y, ORF1a:P2046L, ORF1a:P2685T, ORF1a:T770I, ORF1b:Y710C, S:D1184Y, S:D405N, S:Q954H | M:S4F, ORF1a:P926H, ORF1a:R3164H, S:E516Q, S:K356T |
| IL | 2/28/23 | OQ609210 | 13 | ORF1a:H134Y, ORF1a:P2046L, ORF1a:P2685T, ORF1a:T770I, ORF1b:Y710C, S:D1184Y, S:D405N, S:Q954H | M:S4F, ORF1a:P926H, ORF1a:R3164H, S:E516Q, S:K356T |
| CA | 1/26/23 | PQ017120 | 15 | N:F53Y, ORF1b:R1430K, ORF9b:S50T, S:D405N, S:Q954H | M:S4F, ORF1a:L3027F, ORF1a:P926H, ORF1a:R3164H, S:E516Q, S:F157L, S:G142D, S:K147E, S:K356T, S:W152R |

